# Striatal Dopamine and Skeletal Muscle Energy Metabolism in Older Adults

**DOI:** 10.1101/2025.06.12.25329490

**Authors:** Caterina Rosano, Nico I. Bohnen, Brian LoPresti, Lana M. Chahine, Haley N. Barnes, Stephanie L Studenski, Nancy W. Glynn, Anne B. Newman, David J. Marcinek, Russell T. Hepple, Paul Coen

## Abstract

Dopamine (DA) in the central nervous system is considered a master regulator of mobility performance and vigor, but its mechanistic relationship with skeletal muscle energetics is unclear. We tested the cross-sectional association of striatal DA and skeletal muscle mitochondrial function in 146 older adults participating in the Study of Muscle, Mobility and Aging (75.4 years old, 54% women). Striatal DA was measured using (+)-a-[^11^C] dihydrotetrabenazine (DTBZ) PET imaging for the limbic, sensorimotor, and executive control subregions. Mitochondrial capacity to produce ATP (ATP_max_, mM ATP/s) was measured in vivo using ^31^P magnetic resonance spectroscopy after repeated voluntary muscle contractions. Ex*-vivo* respirometry assays from biopsies of resting muscle captured complementary aspects of mitochondrial function under optimal conditions. In multivariable linear regression models, [^11^C]DTBZ in the limbic striatum, but not other subregions, was positively associated with greater ATPmax *in vivo*, independent of demographics, muscle volume, leg power, white matter hyperintensities, gray matter atrophy, moderate-to-vigorous physical activity and diabetes (β = 0.275, standard error 0.108, p=0.019). [^11^C]DTBZ was not associated with the ex-vivo mitochondrial respiration markers (p>0.2). The role of striatal limbic DA and the energetic capacity of skeletal muscles should be further investigated in older adults.

## INTRODUCTION

Dopaminergic neurotransmission (DA) is essential for successful goal-directed motor behaviors, not only because it regulates decision making and movement coordination, but also because it influences the overall energetic investment and vigor of our actions^1^. Higher DA in the striatum promotes sustained motor performance even in the presence of physiological indicators of fatigue^2–4^, and influences higher energy expenditure^5^. We have shown that higher functional connectivity in the limbic striatal network—a DA-regulated network involved in motivation—is associated with with lower perception of fatigue,^6^ a strong predictor of motor performance. The DA synthetic capacity of the nucleus accumbens (part of the limbic striatum), measured using positron emission tomography, has also been related to vigor in healthy adults^7^. Although it is assumed that striatal DA exerts these effects via the skeletal muscle system, the underlying mechanisms are not entirely clear.

A role for lower DA in frailty and muscle aging has been suggested^8–10^ but it has not been directly tested. Given the influence of DA in the central nervous system on mitochondrial function^11^, a similar effect on the skeletal muscle would be a plausible hypothesis. Reports of mitochondrial function in skeletal muscle in persons with PD have been documented^12, 13^; in vivo measures of muscle metabolism appeared lower in PD compared to controls, although ex-vivo measures were only lower for subgroups of PD. Overall, prior studies did not measure levels of central DA concurrently with skeletal muscle energetics, and were focused on clinical populations. Investigating the dopamine-muscle link in community-dwelling older adults may provide new insights into the role of the DA system in both normal and pathological motor function, particularly in aging populations for whom DA cell loss and muscle function decline frequently co-occur.

The skeletal muscle is a highly plastic tissue that adapts dynamically to neuromuscular signaling, modulating energy metabolism, tone, and strength in response to signals from the central nervous system. The conceptual framework of skeletal muscle health has evolved from a myocentric viewpoint to an integrated perspective that acknowledges the central nervous system’s essential role in modulating muscle contraction and function^10^. Among many muscle features, the capacity to generate energy is a key indicator of muscle function and quality. Mitochondrial dysfunction is implicated in myriad changes in muscle quality, including as a primary driver of aging phenotypes in skeletal muscle^14^. We recently showed the association between mitochondrial function and mobility in older adults may be mediated through muscle contractile properties, such as power and strength^15^, underscoring the primary role of mitochondrial function in influencing performance. However, that work did not account for central nervous system factors.

Magnetic resonance spectroscopy (MRS) represents a noninvasive technique for quantifying in vivo cellular energy metabolism. Phosphorus-31 (^31^P) MRS assesses mitochondrial ATP production. The rate of phosphocreatine (PCr) resynthesis following repeated muscle contractions is used to compute the maximal mitochondrial ATP production rate (ATPmax). Using this methodology, we found ATP max was lower in persons with PD compared to controls^12^. However, in that study we did not measure striatal DA. Reductions in ATPmax within skeletal muscle have been shown with older age, but associations with striatal DA have not been tested.

To address this gap in knowledge, we test the hypothesis that higher striatal DA predicts higher mitochondrial function in the skeletal muscle in community-dwelling older adults. Given the multifaceted influence of striatal DA on mobility control, we examined three striatal subregions based on their relationships with decision making/executive function, sensorimotor control, and motivation/reward processing. To account for the complex inter-relationships between the central nervous and skeletal muscle systems, as well as health conditions and lifestyle, we leveraged a well characterized cohort Study of Muscle, Mobility and Aging (SOMMA) with *in-vivo* and *ex-vivo* measures of muscle mitochondrial function. Based on our prior findings suggesting a role of the reward network in fatigue^6^, and DA-related deficits in skeletal muscle ATPmax in PD^12^ we primarily focused on the association between limbic striatal DA innervation and measures of ATPmax after contractions. Ex-*vivo* respirometry assays of permeabilized fibers from biopsies of resting muscle were also explored because they capture complementary aspects of maximal mitochondrial respiratory capacity under optimal conditions. Imaging measures of muscle adiposity and fat free volume, neuroimaging indices of brain integrity, and a comprehensive characterization of health-related conditions were considered as potential confounders

## METHODS

### Study Sample

A total of 150 community-dwelling adults aged ≥ 70 years participating in the Study of Muscle, Mobility and Aging (SOMMA, https://sommaonline.ucsf.edu)^16^ at the University of Pittsburgh also participated in the Brain Ancillary study, as previously described^17^. Exclusion criteria for the parent study were: inability to walk one-quarter of a mile or climb a flight of stairs; had BMI≥40 kg/m^2^; had an active malignancy or dementia; or any medical contraindication to biopsy or MRI, and walking limitations, defined as inability to complete a 400 meter walk or speed <0.6m/s when walking 4 meters. Among 439 SOMMA participants at the Pittsburgh site, 285 had a muscle biopsy within the prior 12 months; of these, 150 underwent neuroimaging, 2 were ineligible and the others were not interested. Among 150, a total of 146 participants had both PET neuroimaging and at least one measure of muscle mitochondrial function, hence this represents our analytical sample size. Of note, both MRS-ATP and muscle biopsy were available in 115/146 participants. The interval of time between muscle spectroscopy and neuroimaging on average was 10.1 months. The WIRB-Copernicus Group Institutional Review Board approved the study and all participants gave informed written consent (parent study and ancillary numbers 20180764 and 20110230).

### Structural brain imaging

Structural magnetic resonance (MR) images were collected on a Siemens Biograph mMR PET/MR system capable of simultaneous PET and 3 Tesla (3T) MR image acquisition, as previously described^17^. 3T MR images were acquired simultaneously with PET image acquisition and included T1-weighted magnetization-prepared rapid gradient echo (MPRAGE) and Dixon sequences. Distortion correction, registration, and segmentation was obtained via a combination of FreeSurfer and FSL utilizing the minimum preprocessing pipeline and the Human Connectome Project workbench software (version 1.4.0). White matter hyperintensities were mapped using the T2-FLAIR MRI volume via an automated segmentation method that uses fuzzy connectedness to identify hyperintensities. The main measures of interest for these analyses were gray matter volume from total brain, normalized by intracranial volume, and white matter hyperintensities volumes, normalized by total brain volume.

### Brain PET Imaging

We examined the integrity of nigrostriatal dopaminergic terminals using a well-established PET radioligand targeting the type-2 vesicular monoamine transporter (VMAT2), [^11^C]-a-(+)- dihydrotetrabenazine ([^11^C]DTBZ)^17^. VMAT2 is expressed in presynaptic vesicular membranes and transports monoamine neurotransmitters, including dopamine, from the cytosol to storage vesicles. While not specific to dopamine, nearly all (>95%) striatal VMAT2 is associated with storage vesicles in dopamine terminals,^18,19^. Additionally, relative to other presynaptic dopamine targets (e.g dopamine transporter, aromatic amino-acid decarboxylase), VMAT2 is less affected by pharmacologic or lesion-compensatory regulation. In the context of (1) minimal regulation of VMAT2 by dopamine enhancing drugs or compensatory processes, and (2) absence of neurological diseases like Parkinson’s disease as is the case in this aging population, higher in vivo [^11^C]DTBZ striatal binding in neostriatum (caudate nucleus and putamen) and the limbic striatum (anterior ventral striatum) corresponds to higher levels of storage vesicles in the dopamine terminals originating from the subtantia nigra pars compacta and the ventral tegmental area, respectively, and it suggests greater nigrostriatal dopaminergic integrity and signaling capacity. Therefore, we refer to higher/lower [^11^C] DTBZ binding in PET images to indicate higher/lower dopamine nerve terminal integrity.

For each participant, [^11^C]DTBZ (10-15 mCi) was injected intravenously as a slow bolus (20-30 sec) and list-mode acquisition of PET emission data commenced at the start of radiotracer injection and continued for 60 minutes. PET emission data were reconstructed using filtered backprojection with Fourier rebinning into a dynamic series of 20 frames ranging in duration from 15 sec to 300 sec and attenuation correction of PET emission data was performed.

### Brain Imaging Analyses

PMOD software (Zurich, Switzerland) was used for PET data analyses, including motion correction, PET to MR image registration, segmentation, sampling, and tracer kinetic analyses, as previously described^17^. Volumes of interest for sampling of PET image data were defined on the MPRAGE series using the Imperial College London Clinical Imaging Center atlas, which segments the striatum into its functional subdivisions (CIC atlas): associative (the volume-weighted average of [^11^C]DTBZ activity in the anterior putamen, pre-commissural dorsal caudate, and post-commissural dorsal caudate), limbic (anterior ventral striatum) and sensorimotor (posterior putamen). Quantitation of [^11^C]DTBZ specific binding outcomes (BP_ND_) were determined for each volume of interest using the simplified reference tissue method and occipital cortex as a reference tissue^17^.

### Maximal ATP Production

Maximal mitochondrial ATP production (ATPmax) was quantified using ^31^P magnetic resonance spectroscopy to measure the rate of phosphocreatine regeneration following a short bout of voluntary isometric muscle contractions. The rate of phosphocreatine resynthesis following cessation of muscle contraction is an indicator of the capacity to produce ATP by mitochondrial oxidative phosphorylation in skeletal muscle tissue ^20, 21^. A 3 Tesla MR scanner (Siemens Medical System – Prisma) using a 12” dual-tuned, surface radiofrequency (RF) coil (PulseTeq, Limited) placed over the right distal lateralis was used to collect ^31^P spectra. Participants performed two bouts (30 and 36 seconds) of isometric knee extension, followed by a 6-minute rest, with the instructions of “kicking as fast and as strong as possible” against the resistance of an ankle strap. Phosphocreatine recovery rate after exercise was fit and the time-constant of the mono-exponential fit was used to calculate ATPmax, as previously described.^19^

### Skeletal Muscle Respiration

The skeletal muscle biopsy was taken from the medial vastus lateralis after a 12-hr fast and limited exercise for 48 hours prior to the procedure.^13^ A portion (∼20mg) of the specimen was placed in a biopsy preserving solution for high-resolution respirometry, as previously described.^22^ Myofiber bundles (∼2-3mg) were prepared by gently separating the fibers using fine-point tweezers and chemically permeabilized in a solution containing saponin. After weighing, the permeabilized myofiber bundles were placed into Oxygraph-2K respirometer chambers (O2K, Oroboros Instruments, Austria). The respirometry protocol was performed in duplicate for each muscle biopsy specimen. Complex I supported leak respiration (LEAK) was elicited by addition of pyruvate (5mM) and malate (2mM) to the chamber. Maximal Complex I supported OXPHOS (Max CI OXPHOS) was achieved after addition of ADP (4.2mM) and Glutamate (10 mM) and maximal complex I- and II-supported oxidative phosphorylation (Max OXPHOS) was measured following the addition of succinate (10mM). Finally, maximal electron transport system capacity (Max ETS) was assessed following an FCCP titer (up to 2mM). Assays were run at 37°C within a specific range of O_2_ concentration (400-200μm). Steady-state oxygen flux was normalized to the fiber bundle wet weight using Datlab 7.4 software. All were measured in pmol/(s*mg). Technician was controlled for in analyses.

### Other Assessments of Muscle Integrity

*Leg Power Measurement.* Knee extensor leg power was assessed using a Keiser Air 420 exercise machine. The right leg was used unless contraindicated because of prior joint replacement or range of motion restrictions. Anyone with a recent (6 months) stroke, aneurysm, cerebral hemorrhage, or systolic blood pressure >180 or <90 mmHg was excluded from testing. If the examiner observed or the participant reported excessive pain or discomfort on a standard scale, testing was discontinued. Resistance to test power was based on determination of the 1 repetition maximum leg extensor strength. Power was tested at 40%, 50%, 60%, and 70% of the participant’s maximum^23^.

*Muscle tissue composition.* An MRI scan was taken of the whole body to assess body composition. All MRI data were processed by the AMRA Company. Images were analyzed using AMRA Researcher (AMRA Medical AB). Briefly, the image analysis consisted of image calibration, fusion of image stacks, image segmentation, and quantification of fat and muscle volumes^24, 25^ and included manual quality control by a blinded trained operator. Among the composition parameters, there were two who are of interest for these analyses: fat-free muscle volume and muscle fat infiltration, computed as the mean fat as a fraction of the fat-free muscle volume of the right and left anterior and posterior thigh.

### Health-related variables

Information on age in years was recorded at the time of baseline brain MRI, the other variables were from the baseline visit of the parent SOMMA.

*Non-Hispanic white vs. other racial or ethnic minority* was by self-report. *Cognitive function was assessed using* the Montreal Cognitive assessment (MoCA), with a range from 0-30. *Multimorbidity* was based on number of conditions out of a total of 11, and it was assessed via self-reported physician diagnosis (cancer, excluding nonmelanoma skin cancer, cardiac arrhythmia, chronic kidney disease, chronic obstructive pulmonary disease, coronary heart disease, congestive heart failure, dementia, diabetes, stroke, aortic stenosis, and depression). Depression was assessed via the Center of Epidemiologic Studies of Depression-Scale. *Diabetes* was assessed by self-report, use of hypoglycemic meds, or HbA1c =6.5%. *Hypertension* was assessed based on systolic blood pressure ≥ 140 mmHg. *Prescription medications* in past 30 days were counted, including anti-dopaminergic, anticholinergic, and central nervous system active medications. Use of anti-depressants was assessed separately, and included Selective Serotonin Reuptake Inhibitors, non-selective monoamine reuptake inhibitors, Monoamine Oxidase Inhibitors. *Body mass index (BMI, kg/m*^2^) was calculated from height (Harpenden stadiometer, Dyved, UK) and weight (balance beam or digital scale), measured without shoes and with light clothing. *Physical activity* was quantified with wrist-worn accelerometry using the GGIR R package as average time spent in moderate-to-vigorous physical activity (MVPA, min/day)^26^.

### Statistical methods

Baseline characteristics were shown as n (%) or mean +/- standard deviation. Distributions of the main variables of interest were examined to determine whether transformation were required. A log-transformation was applied to non-normally distributed outcome measures. Outcome measures were standardized to interpret results across different units of measurement. Multi-collinearity of covariates was addressed via examination of variance inflation factors and condition indices. This was followed, as needed, by parsimonious selection of the best predictors within classes of predictors and covariates.

A set of multivariable linear regression models tested the hypothesis that higher [^11^C]DTBZ in striatal subregions was associated with higher ATPmax; models were first adjusted for demographics and subsequently for potential confounders, including health-related variables, muscle measures (ex-vivo mitochondrial function, power, adiposity), and brain measures (cognition, neuroimaging).

Each potential confounder entered separate age- and sex-adjusted models with ATPmax or striatal DA as the outcome. Given our interest on muscle mitochondrial function, we explored the association between [^11^C]DTBZ in striatal subregions and ex-vivo muscle mitochondrial function, adjusted for age, sex and technician. Results were expressed as beta coefficients (β) with standard error (SE). Analyses were completed in SAS version 9.4 using data as of November 2024.

## RESULTS

Participants of the SOMMA-Brain study were on average 75 years old, about half were women, and the majority was non-Hispanic white. The average cognitive score, prevalence of diabetes and hypertension indicate this sample is overall well-functioning (Table 1).

**Table 1.**
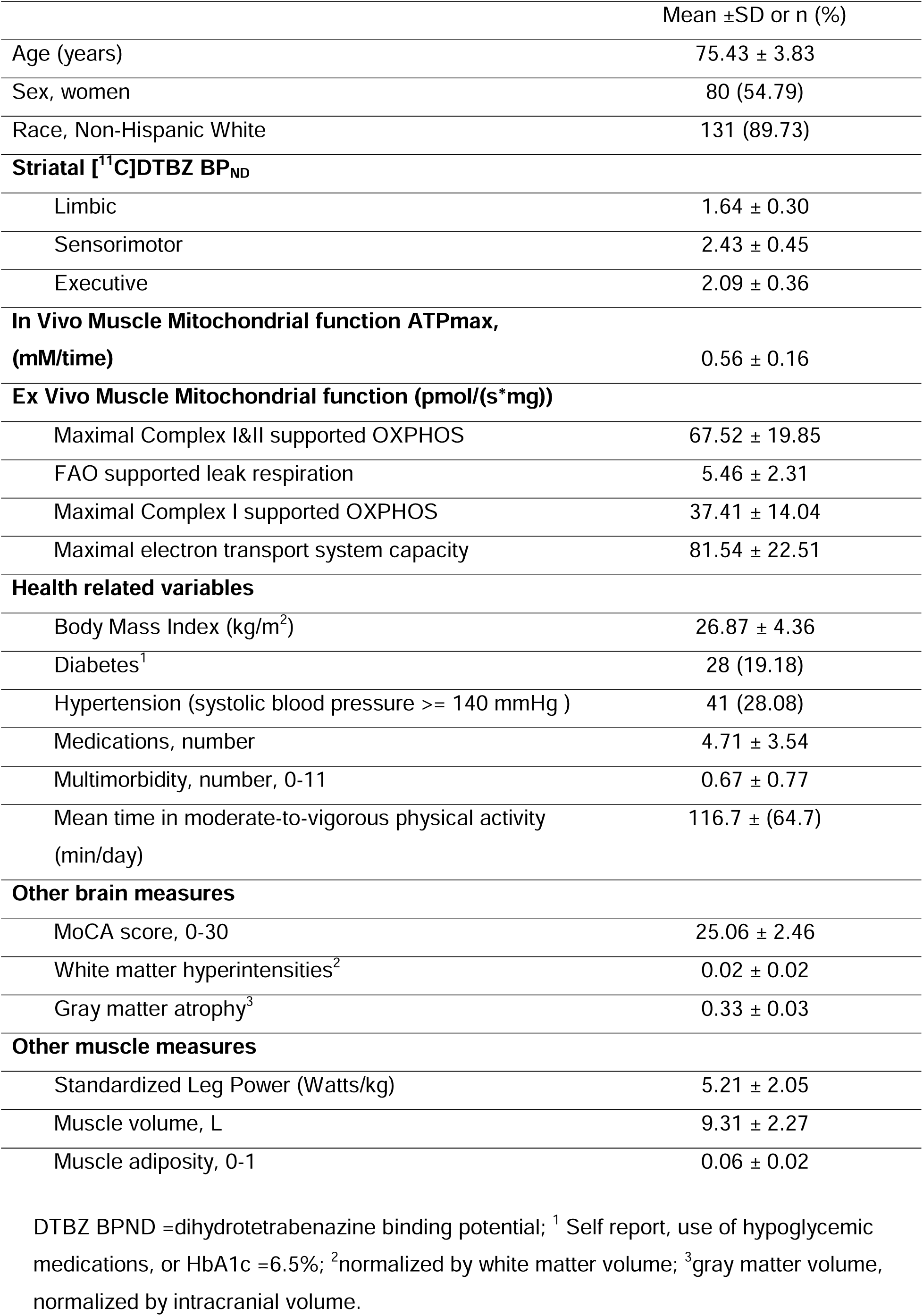
Study Sample Characteristics in 146 participants: Study of Muscle, Mobility and Aging Brain Ancillary Study.

When investigating potential confounders, we found ATPmax was significantly associated with all four ex-vivo measures of mitochondrial function (p≤0.008), as well as with diabetes, leg power, and MVPA (age and sex adjusted b [standard error], p value: -0.60 [0.32], p=0.007; 0.31 [0.11], p=0.015; and 0.35 [0.11] p=0.005, respectively, see Supplemental Table 1). Higher [^11^C]DTBZ in the limbic striatum was not associated with any of the potential confounders (p>0.2 for all).

Higher [^11^C]DTBZ in the limbic striatum correlated with ATPmax, independent of age and sex (Table 2, Model 1). Further adjustment for diabetes, leg power or MVPA did not modify the results (Table 2, Model 2). Adding other markers of structural integrity of muscle or brain to the models did not substantially modify the main association of [^11^C]DTBZ binding with ATP max (Table 2).

**Table 2.** Association of [11]C DTBZ PET binding in the limbic striatal region with ATPmax in skeletal muscle.

|  | Model 1 |  | Model 2 |  | Model 3 |  |
| --- | --- | --- | --- | --- | --- | --- |
|  | Beta (SE) | *p* | Beta (SE) | *p* | Beta (SE) | *p* |
| Limbic <sup>1</sup> | 0.267<br>(0.118) | 0.034 | 0.263<br>(0.112) | 0.029 | 0.275 (0.108) | 0.019 |
| Sensorimotor <sup>2</sup> | 0.142<br>(0.113) | 0.218 |  |  |  |  |
| Associative <sup>3</sup> | 0.172<br>(0.112) | 0.137 |  |  |  |  |

1: Unit: 0.2861, 2: 0.4534, 3:0.3670
Model 1: adjusted for age at scan, sex
Model 2: further adjusted for diabetes, moderate-to-vigorous physical activity, muscle power,
Model 3: further adjusted for muscle volume, brain atrophy, small vessel disease

Associations of sensorimotor or associative striatum with ATPmax were not statistically significant (Table 2), hence were not further investigated. Exploratory analyses of [^11^C]DTBZ binding with *ex-vivo* mitochondrial markers did not yield statistically significant results (p>0.186, Supplemental Table 2).

## DISCUSSION

We found a positive and significant correlation between DA measured via PET in the limbic striatum and greater mitochondrial capacity for ATP production in skeletal muscle after repeated contractions. This association was robust to adjustment for measures of brain and skeletal muscle integrity, including brain atrophy and small vessel diseases, muscle volume and power. These results have relevance for potential new strategies to promote muscle energetic capacity in older age, especially considering recent evidence that enhancing DA neurotransmission promotes gait speed in slow walking older adults^27^. Our study adds to the literature by including assessments of both striatal DA and muscle mitochondrial function, as well as comprehensive characterization of brain and muscle integrity and health-related conditions. The association of DA in the central nervous system with skeletal muscle mitochondrial function has not been directly tested in humans. Studies in the context of neurological diseases, in particular PD, support a link between DA in the central nervous system and processes indirectly related to energy metabolism in the skeletal muscle, such as synchronization of electrical activity^28^, and blood flow and metabolism^29^. Reductions in ATPmax within skeletal muscle have been shown both in aging and in individuals with PD^10, 12^. However, these studies did not have concurrent measures of both central DA levels and muscle mitochondrial function.

The link between central DA and muscle energetics is biologically plausible. DA modulates activity of neuronal efferents to the spinal cord and from there to skeletal muscles. These supra-spinal processes can influence sustained muscle contractions and physical performance, potentially inducing neuromuscular junction remodeling, which in turn promotes muscle mitochondrial function^22^. In this context, it is interesting to note the null results of this study. Maximal muscle mitochondrial respiratory capacity measured ex vivo in permeabilized muscle fiber bundles was not significantly associated with striatal DA; this finding was somewhat surprising, given the association of ATPmax in the same muscle group. A potential explanation for this null finding is that the two analytical approaches employed here (respirometry and ^31^P MRS) measure different features of mitochondrial function and under different conditions. The ex-vivo respirometry protocol assessed the maximal capacity of the skeletal muscle fibers to consume oxygen under optimal concentrations of substrate, ADP and oxygen. As such, measurement of respiratory capacity is therefore independent of contraction, blood flow, and especially independent of neural input. Conversely, assessment of ATPmax via ^31^P MRS is an integrated system measure that calculates ATP production under physiological blood flow, neural input, and cellular processes, while the ex vivo respirometry measures report oxygen consumption in isolated muscle fibers, as a proxy for oxidative phosphorylation capacity under optimal conditions. If there is reduced coupling of oxygen consumption to ATP production (lower generation of ATP for a given rate of mitochondrial oxygen consumption), then it could manifest in lower ATPmax without influencing the ex vivo Max OXPHOS measure^30^. Our results are consistent with those of a recent study of ex-vivo mitochondrial respirometry capacity in persons with and without PD,^13^ which found no between-group differences in CI levels. Of note, this study also found a subset of PD displayed deficits in mitochondrial respiratory Complex I but not in Complex II-IV^13^, thus underscores the importance of further examining the link between DA and in-depth measures of mitochondrial function for subgroups of individuals with lower mobility.

We found associations were stronger for the limbic striatum compared to other striatal subregions, consistent with prior work showing an association of greater integrity in this subregion with less fatigue^6^ and more vigor^31^. While we did not assess the mechanisms of this distinct striatal spatial distribution, we can advance several potential explanations. One explanation for the differential association of the limbic, but not other striatal subregions, with ATPmax, may be because of differences in the midbrain DA neurons innervating these striatal subregions. The radioligand used in this study captures DA content in presynaptic vesicles whose axons and neurons originate in different regions of the midbrain; in other words, the PET signal in the striatum is primarily reflecting the integrity of DA neurons in the midbrain. The DA innervation of the limbic striatum originates in the ventral tegmental area (VTA), whereas the innervation of the other striatal subregions originates in the substantia nigra (SN) pars compacta. It has been shown that efferent projections from the VTA, but not the SN, influence the autonomic control of the cardiovascular system, resulting in skeletal muscle vasodilatation. Our results raise the possibility that, compared to the SN, the VTA may be preferentially involved in skeletal muscle vascular response to muscle contractions and supply of substrate and oxygen needed for ATP production^32^. This pathway may also explain why we did not observe an association with ex-vivo measures of mitochondrial function, which are independent of blood flow. Although other striatal subregions, and in particular the sensorimotor striatum, are engaged in motor control processes, the limbic striatum is less affected by aging compared to other striatal subregions^33^, thus it may play a larger compensatory role in regulating the complex aspects of motor control. A ventro-dorsal gradient of compensation has been observed in animal models, with the limbic (ventral) striatum compensating for age-related neuronal loss in the dorsal (sensorimotor) striatum. An effect on muscle tone has been shown specifically after stimulating the limbic but not the sensorimotor striatum in animal models^34^. However, in the present study, the association of limbic striatum with muscle power was not statistically significant. Prior studies assessing the link of DA with muscle in non-PD older adults yield mixed results. One study using acute carbidopa/levodopa challenge to increase central DA levels, found an increase in motor performance but not in grip strength^8^. In two separate studies using the DA genotype catechol-O-methyltransferase as a surrogate of central DA levels, we did not find statistically significant associations with muscle strength^35, 36^. Future studies should focus on investigating whether the DA-ATPmax association would translate to higher muscle power or strength.

The results of this study should be interpreted in the context of its limitations. First, given the cross-sectional nature of our analyses, we cannot exclude that limbic DA may not be the driver but the result of muscle mitochondrial function or that there is a bidirectional interaction. It has been shown that brain-muscle relationships can be driven by skeletal muscle, both via neural and non-neural/humoral pathways. Non-neural, bidirectional pathways linking central DA and skeletal muscle metabolism via circulating endocrine signals and growth factors^37, 38^ have been shown. A direct effect of DA on mitochondrial function in skeletal muscles has been shown in vitro in animal models^39^. Although this possibility would not explain the distinct association with the limbic region, it is nonetheless an intriguing line of research that deserves further study. Secondly, we tested three striatal subregions in a relatively small sample; a Sidak correction for multiple comparisons would have adjusted the significance threshold to 0.0169, rendering our finding (age and sex-adjusted p=0.03) non-significant. However, due to the novelty of the study, we argue that a more lenient threshold of p<0.05 is justified. Moreover, we had an a priori hypothesis pointing at the limbic striatum as the primary independent variable and ATP max as the outcome^6, 12^. Lastly, we note the association between striatal DA and ATPmax remained significant even after adjustment for several covariates, thus providing confidence in this result. Another limitation is that the DA and muscle measures were not taken on the same day. However, relatively short-term changes in mitochondrial function or DA neurotransmission in the absence of clinically overt diseases are unlikely, and adjustment for the interval of time between brain scan and muscle biopsy did not modify the results.

In conclusion, this study begins to explore potential relationships between striatal DA and skeletal muscle energetics in older adults. To our knowledge, this is the first study suggesting a DA-muscle energetics connection in older adults free of clinical neurological diagnoses. While this investigation was not designed to establish a mechanistic link between central DA and muscle energetic processes, it provides novel information that warrants future study.

DTBZ BPND =dihydrotetrabenazine binding potential; ^1^ Self report, use of hypoglycemic medications, or HbA1c =6.5%; ^2^normalized by white matter volume; ^3^gray matter volume, normalized by intracranial volume.

## Data Availability

SOMMA data (November 2024 release) are publicly available by request at https://sommaonline.ucsf.edu/.

## Funding Information

The Study of Muscle, Mobility and Aging is supported by funding from the National Institute on Aging, **U01AG061393, R01AG075025, AG059416**. Study infrastructure support was funded in part by NIA Claude D. Pepper Older American Independence Centers at University of Pittsburgh (**P30AG024827**) and Wake Forest University (**P30AG021332**) and the Clinical and Translational Science Institutes, funded by the National Center for Advancing Translational Science, at Wake Forest University (**UL1 0TR001420**)

**Permission Statement:** Not Applicable

## Author Contributions

Ms. Barnes and Drs. Rosano and Coen had full access to all of the data for the study and take responsibility for the integrity of the data and accuracy of the data analyses. All authors: interpretation of data, critical revision of manuscript for important intellectual content. All authors read and approved the submitted manuscript.

## Acknowledgements

We acknowledge all the staff and investigators, and we appreciate all the participants.

---

**Supplemental Table 1.**
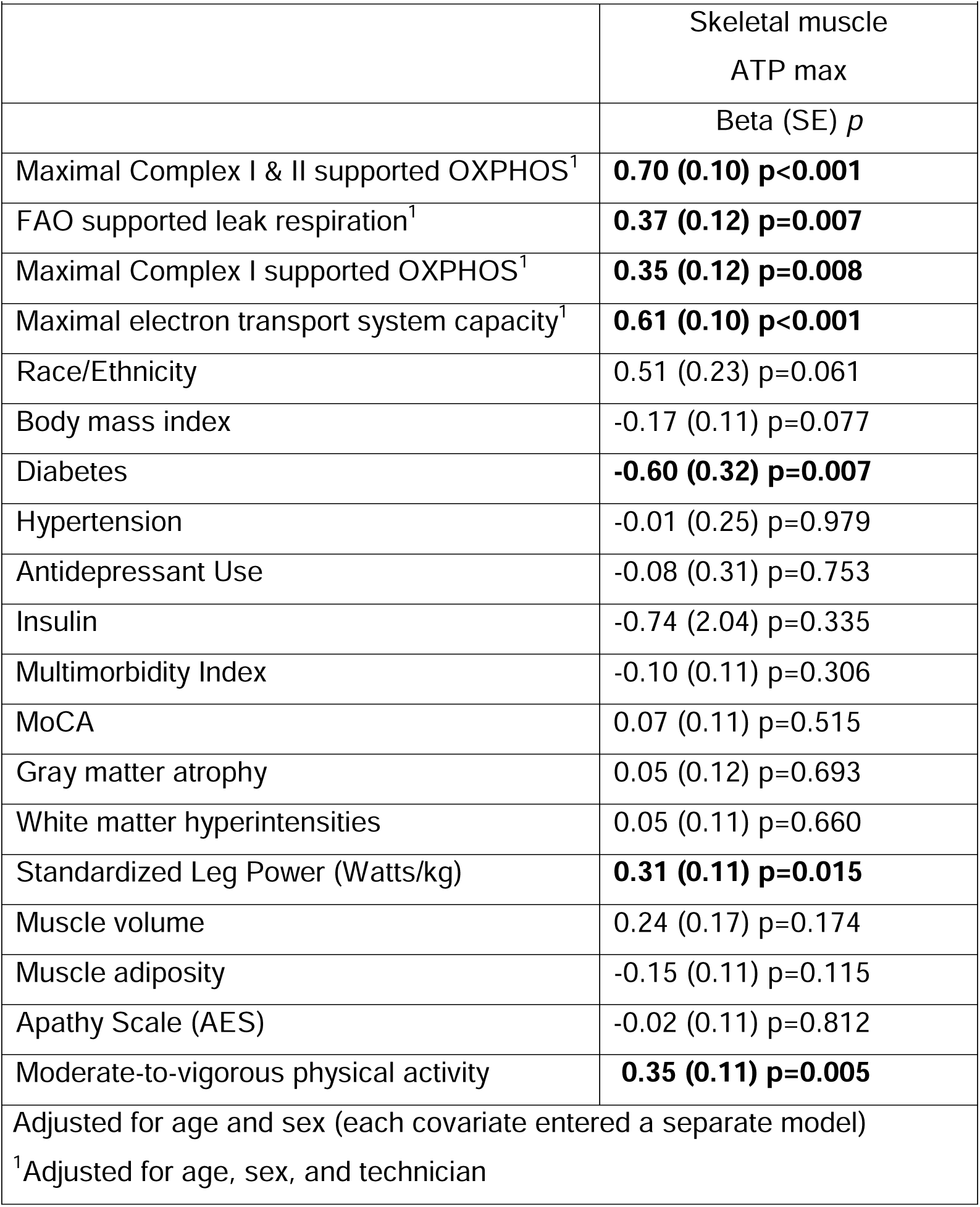
Associations of confounder covariates with the main outcome.

|  |  |
| --- | --- |
|  | Skeletal muscle<br>ATP max |
|  | Beta (SE) *p* |
| Maximal Complex I & II supported OXPHOS<sup>1</sup> | **0.70 (0.10) p<0.001** |
| FAO supported leak respiration<sup>1</sup> | **0.37 (0.12) p=0.007** |
| Maximal Complex I supported OXPHOS<sup>1</sup> | **0.35 (0.12) p=0.008** |
| Maximal electron transport system capacity<sup>1</sup> | **0.61 (0.10) p<0.001** |
| Race/Ethnicity | 0.51 (0.23) p=0.061 |
| Body mass index | -0.17 (0.11) p=0.077 |
| Diabetes | **-0.60 (0.32) p=0.007** |
| Hypertension | -0.01 (0.25) p=0.979 |
| Antidepressant Use | -0.08 (0.31) p=0.753 |
| Insulin | -0.74 (2.04) p=0.335 |
| Multimorbidity Index | -0.10 (0.11) p=0.306 |
| MoCA | 0.07 (0.11) p=0.515 |
| Gray matter atrophy | 0.05 (0.12) p=0.693 |
| White matter hyperintensities | 0.05 (0.11) p=0.660 |
| Standardized Leg Power (Watts/kg) | **0.31 (0.11) p=0.015** |
| Muscle volume | 0.24 (0.17) p=0.174 |
| Muscle adiposity | -0.15 (0.11) p=0.115 |
| Apathy Scale (AES) | -0.02 (0.11) p=0.812 |
| Moderate-to-vigorous physical activity | **0.35 (0.11) p=0.005** |
| Adjusted for age and sex (each covariate entered a separate model) |  |
| <sup>1</sup>Adjusted for age, sex, and technician |  |

**Supplemental Table 2.** Regression coefficients of Dopamine in the Limbic striatum with ex-vivo markers of skeletal muscle mitochondrial function.

|  | B (SE)<sup>1</sup> | *p* |
| --- | --- | --- |
| Maximal complex I- and II-supported oxidative phosphorylation (Max OXPHOS) | 0.10 (0.10) | 0.337 |
| Complex I supported leak respiration (LEAK) was elicited by addition of pyruvate (5mM) and malate (2mM) to the chamber | 0.10 (0.10) | 0.289 |
| Maximal Complex I supported OXPHOS (Max CI OXPHOS) | 0.13 (0.10) | 0.186 |
| Maximal electron transport system capacity (Max ETS) was assessed following an FCCP titer (up to 2mM) | 0.13 (0.10) | 0.209 |
| Unit: 0.2951. <sup>1</sup> Adjusted for age, sex, and technician |  |  |

